# High-Throughput Observational Evidence Generation Using Linked Electronic Health Record and Claims Data

**DOI:** 10.64898/2026.04.07.26350300

**Authors:** Jeremy Coyle, Neil Shah, Alan Hubbard, Arjun Mukerji, Miriam Chappelka, Neil Sanghavi, Saurabh Gombar

## Abstract

**Background:** Many consequential treatment decisions involve off-label or head-to-head choices in complex, comorbid populations routinely excluded from randomized trials, or other prospective analyses. For these decisions, comparative evidence is often entirely absent. Although observational data include these patients, findings are difficult to synthesize because studies differ in cohort definitions, confounder measurement, follow-up periods, and reported outcomes. Prior systems for large scale evidence generation have largely stopped at data preparation, limiting the usefulness of their outputs to decision makers.

**Methods:** We developed a high-throughput evidence-generation workflow using linked EHR and claims data. A prespecified causal-measurement architecture was applied consistently across scenarios, including three post-index follow-up windows through two years; 28 comorbidities; 14 healthcare resource utilization categories; 30 laboratory measures with 57 binary thresholds; 43 adverse-event categories; and evaluated 1038 clinically important outcomes. Scalable collaborative targeted learning (C-TMLE) generated confounding-adjusted, actionable treatment comparisons, with unadjusted estimates reported for transparency.

**Results:** Across 135 clinical scenarios, the workflow generated 210,584,509 outcome evaluations. Each evaluation represented an outcome, follow-up window, treatment contrast, population stratum, and estimator, accompanied by diagnostic information. These results were synthesized into 7,500 narrative summaries and underwent structured clinical and statistical quality control.

**Conclusions:** Standardized, high-throughput workflows can move evidence generation beyond fragmented individual studies toward comprehensive evidence packages. By making treatment-effect heterogeneity visible across clinically meaningful subgroups, this shared evidence base can support precision medicine and reduce redundant stakeholder-specific studies.

## Introduction

The clinical decisions with the least evidence are often the most consequential: which therapy to choose for a patient whose comorbidities no trial enrolled, whether an off-label drug is safe in a comorbid population, or how benefits and harms differ across subgroups trials cannot resolve.

The problem is not merely the absence of a randomized trial but of any comparative evidence: the relevant patients are routinely excluded, 77% of trials exclude more than half of patients with the disease of interest based on patient history ^1^, and the relevant head-to-head comparisons are seldom run. These patients still need evidence-based decisions. The absence of trial data does not remove the need, only shifts it to real-world data.

Real-world evidence is now central to clinical, payer, and regulatory decisions. Yet the literature remains hard to synthesize: groups address identical questions using divergent design and analytic conventions, making real heterogeneity hard to distinguish from methodological noise ^2,3^. Two issues drive this fragmentation. First, the same question can be operationalized in many ways: index date, censoring, and follow-up choices each move the estimate ^4^. Second, outcomes are usually chosen through a narrow specialty lens, so a cardiologist’s endpoint may be irrelevant to a payer or patient ^5,6^.

Closing this gap requires four elements: (1) data built for causal analysis (active-comparator, new-user cohorts with correctly-timed confounder and outcome capture); (2) a comprehensive measurement framework spanning the outcomes different stakeholders need; (3) rigorous, robust statistical methods; and (4) systematic execution at scale with transparent dissemination countering publication bias and analytic opportunism ^7,8^. When all groups pre-specify the same outcomes and windows before results are known, the evidence becomes a shared resource: clinicians receive clinical endpoints, payers utilization data, and life-science companies subgroup efficacy and safety profiles. Generating observational evidence at scale is not new: the OHDSI LEGEND framework estimates treatment effects across many exposures and outcomes with empirical calibration ^9,10^, the FDA Sentinel System conducts distributed post-market safety surveillance ^11^, and TWOSIDES catalogued drug–effect associations by mining spontaneous adverse-event reports ^12^. We build on that direction. Here we describe such a workflow applied to linked EHR and claims data across 135 clinical scenarios, combining rigorous measurement, transparent statistical reporting, and structured quality control.

## Methods

The workflow (Figure 1) generates rigorous observational evidence where trials are absent, complementing rather than replacing them.

**Figure 1.**
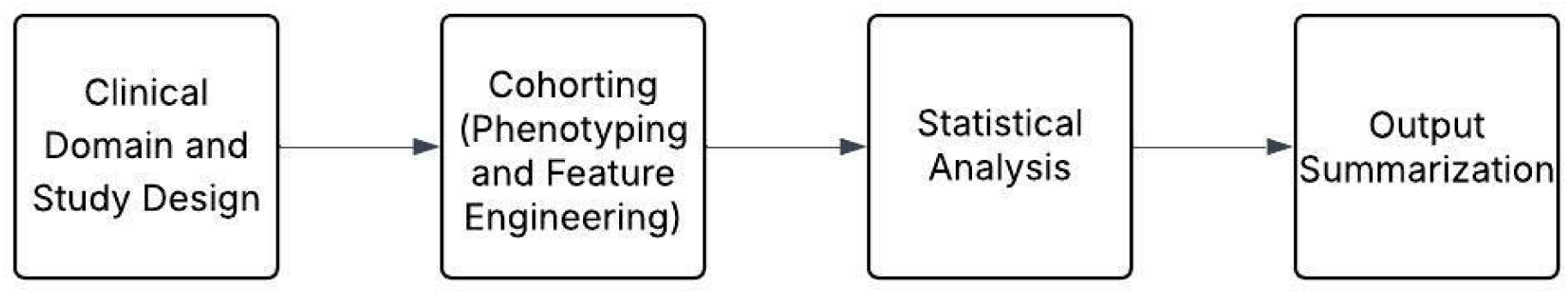
Overall Process Flow. Schematic of the high-throughput evidence-generation pipeline: clinicians select a clinical domain and prespecify the study design; linked electronic health record (EHR) and claims data are then transformed into clinician-reviewed cohorts and baseline features through phenotyping and feature engineering. Outputs include standardized multi-outcome effect estimates with uncertainty, prespecified subgroup and sensitivity analyses, and narrative summaries to support transparent, reproducible reuse.

### Study Design and Data

We conducted a high-throughput observational study on linked EHR and administrative claims data from the Atropos Apollo source database of 142 million patients, part of the <u>Atropos</u> <u>Evidence Network</u>, with integrated demographics, diagnoses, procedures, labs, vitals, provider, and payer-cost data. The 142-million figure is the source database, not any analytic cohort. Across the portfolio, analytic cohorts (summed across arms) ranged from several hundred to ∼14,500 patients after downsampling, each arm capped at 5,000 by uniform random sampling (a precision cost, not a bias; estimates target the sampled population and are not reweighted). Index dates spanned 2000 onward, concentrated in recent years by treatment availability.

Because longitudinal records extend for years pre-index, baseline measurements span both ICD-9 and (post-2015) ICD-10 eras, both mapped. Reporting follows the RECORD ^13^ extension of the STROBE guideline ^14^.

### Clinical Scenario Selection and Cohort Construction

Clinicians selected clinical domains (e.g., cardiovascular, mental health, rheumatology) and, within them, 135 treatment-comparison scenarios. We used an active-comparator, new-user design following target-trial-emulation principles ^15^: the active comparator reduces confounding by indication, the new-user requirement avoids prevalent-user and immortal-time biases. The index date, baseline and follow-up windows, enrollment minimums (≥3 months pre-index, ≥6 months post-index), and censoring rule are summarized in Figure S1 and specified in full in Supplementary Information S1, Section 1. Because follow-up is censored at the earliest of death, disenrollment, treatment discontinuation, or switch to a second-line agent, estimates are as-treated contrasts conditioned on remaining on index treatment. This template is applied uniformly across scenarios. Each scenario’s full protocol is prespecified and delivered in its evidence report rather than enumerated here.

#### Measurement Framework

The measurement architecture is this work’s central methodological contribution. Rather than choosing outcomes to match a single specialty or stakeholder, we pre-specified a standardized set of measurements and applied it uniformly across all 135 scenarios (Table 1), so one design serves patients, clinicians, payers, policymakers, and life-science companies without any commissioning a separate study. All windows are anchored to the index date: baseline characteristics in a single 365-to-15-day pre-index window (a 15-day washout prevents peri-prescribing lab draws from contaminating baseline), and post-index outcomes across three windows spanning acute through two-year follow-up (Supplementary Information S1, Section 1). These windows capture clinically distinct phases — acute effects and early tolerability (0–90 days), intermediate-term efficacy and durability (91–365 days), and longer-term outcomes (366– 730 days) — while limiting the multiplicity that finer or overlapping intervals would introduce.

**Table 1.**
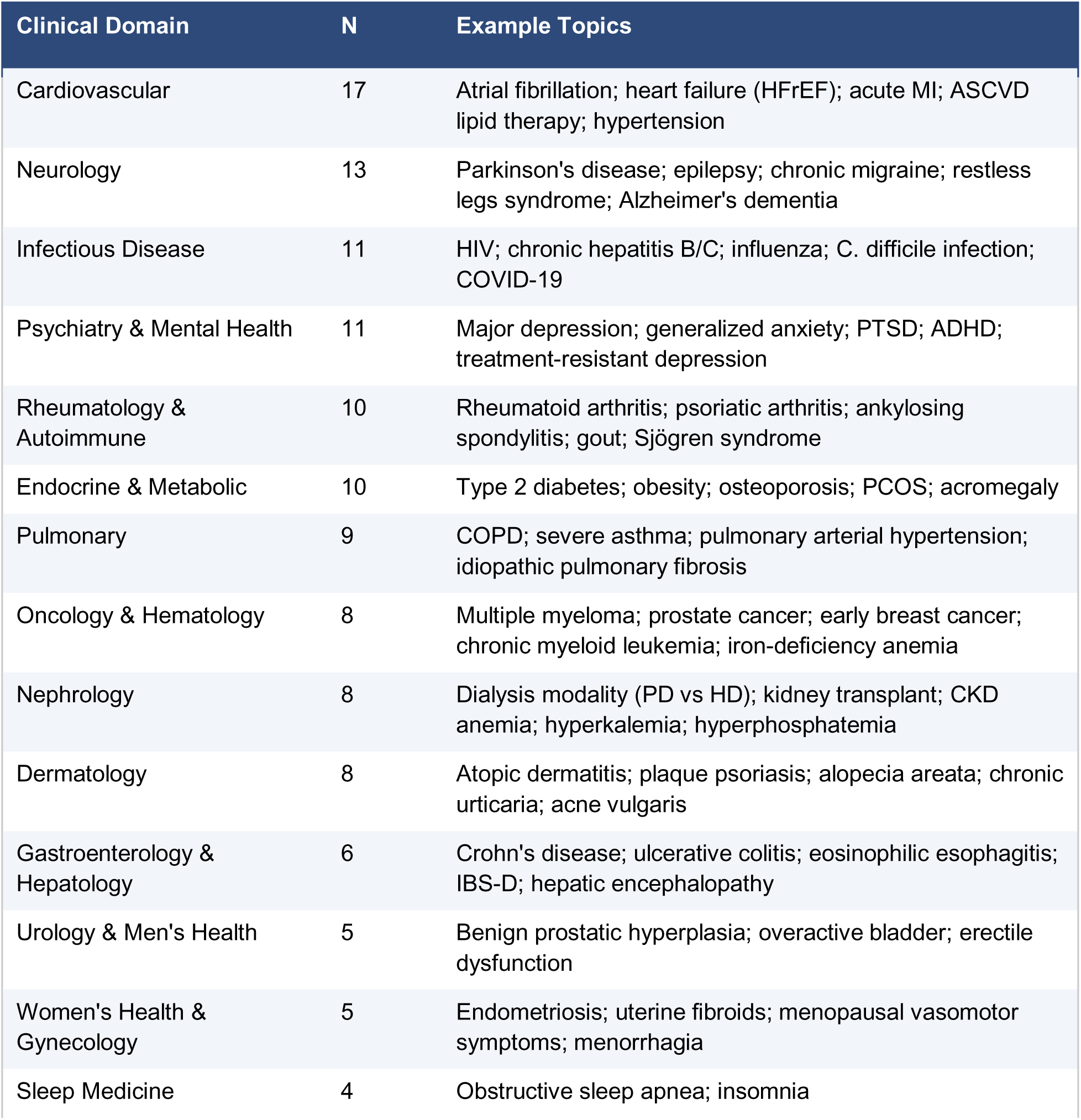
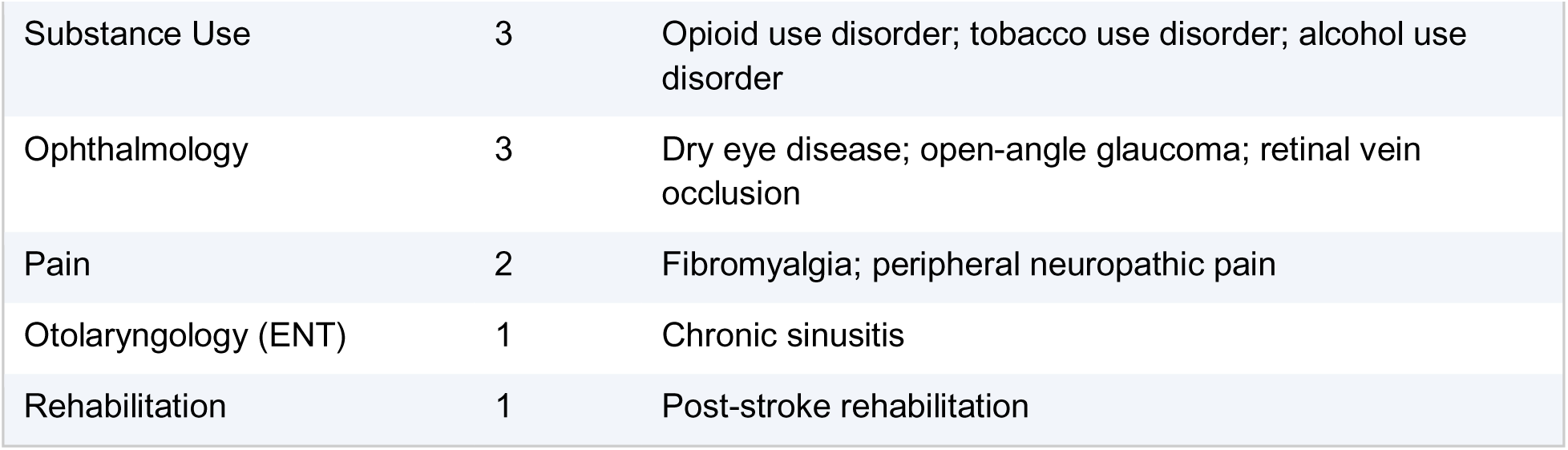
Clinical domains and example topics across the 135 treatment-comparison scenarios. N is the number of scenarios per domain; example topics are illustrative, not exhaustive. The full list of scenarios is provided in Supplementary Information S1, Section 12.

| Clinical Domain | N | Example Topics |
| --- | --- | --- |
| Cardiovascular | 17 | Atrial fibrillation; heart failure (HFrEF); acute MI; ASCVD lipid therapy; hypertension |
| Neurology | 13 | Parkinson's disease; epilepsy; chronic migraine; restless legs syndrome; Alzheimer's dementia |
| Infectious Disease | 11 | HIV; chronic hepatitis B/C; influenza; C. difficile infection; COVID-19 |
| Psychiatry & Mental Health | 11 | Major depression; generalized anxiety; PTSD; ADHD; treatment-resistant depression |
| Rheumatology & Autoimmune | 10 | Rheumatoid arthritis; psoriatic arthritis; ankylosing spondylitis; gout; Sjögren syndrome |
| Endocrine & Metabolic | 10 | Type 2 diabetes; obesity; osteoporosis; PCOS; acromegaly |
| Pulmonary | 9 | COPD; severe asthma; pulmonary arterial hypertension; idiopathic pulmonary fibrosis |
| Oncology & Hematology | 8 | Multiple myeloma; prostate cancer; early breast cancer; chronic myeloid leukemia; iron-deficiency anemia |
| Nephrology | 8 | Dialysis modality (PD vs HD); kidney transplant; CKD anemia; hyperkalemia; hyperphosphatemia |
| Dermatology | 8 | Atopic dermatitis; plaque psoriasis; alopecia areata; chronic urticaria; acne vulgaris |
| Gastroenterology & Hepatology | 6 | Crohn's disease; ulcerative colitis; eosinophilic esophagitis; IBS-D; hepatic encephalopathy |
| Urology & Men's Health | 5 | Benign prostatic hyperplasia; overactive bladder; erectile dysfunction |
| Women's Health & Gynecology | 5 | Endometriosis; uterine fibroids; menopausal vasomotor symptoms; menorrhagia |
| Sleep Medicine | 4 | Obstructive sleep apnea; insomnia |
| Substance Use | 3 | Opioid use disorder; tobacco use disorder; alcohol use disorder |
| Ophthalmology | 3 | Dry eye disease; open-angle glaucoma; retinal vein occlusion |
| Pain | 2 | Fibromyalgia; peripheral neuropathic pain |
| Otolaryngology (ENT) | 1 | Chronic sinusitis |
| Rehabilitation | 1 | Post-stroke rehabilitation |

Applied within each window, the framework spans comorbidities, healthcare-resource utilization, laboratory and vital-sign measures (with change-from-baseline values and guideline-based thresholds), and adverse events (Table 2; ^16–19)^. These candidates serve both confounding adjustment and subgroup screening. Complete variable lists, code sets, plausible ranges, thresholds, and data-quality rules are in Supplementary Information S1, Sections 1–7.

**Table 2.**
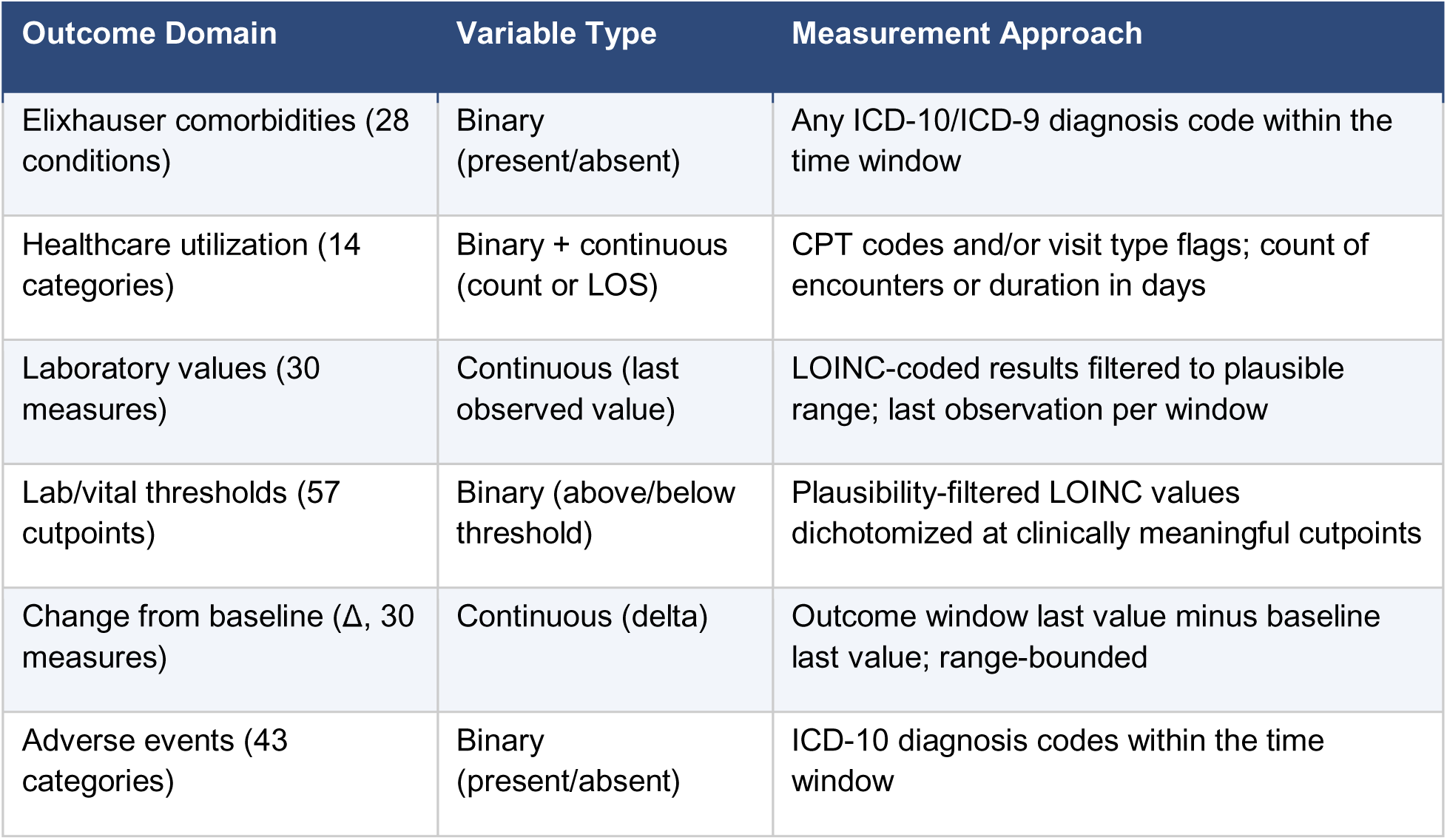
Outcome Domains, Variable Types, and Measurement Approach.

### Statistical Analysis

#### Estimands and Estimators

We chose our statistical strategy to (1) estimate the data-generating distribution without arbitrary modeling assumptions, (2) adapt model complexity to sample size and problem difficulty, (3) target the bias–variance trade-off at the treatment effect, not outcome prediction, (4) yield valid inference under data-adaptive estimation, and (5) rest on estimators with formal theory and prior empirical evaluation. Together, these criteria address the two failure modes that most undermine reproducibility: model misspecification and analyst degrees of freedom. Because the pipeline is pre-specified and runs deterministically in a pinned container image, two analysts obtain the same estimate.

Our primary estimator is a doubly robust collaborative TMLE (C-TMLE) — the scalable variant of Ju et al. ^20^ — adjusting for measured baseline confounding. TMLE combines an outcome regression and a propensity model with a targeting step, giving consistent, semiparametrically efficient treatment-specific means when either nuisance model is consistent. Nuisance parameters use cross-fitted gradient-boosted (XGBoost) learners, chosen for scalability and robustness across this portfolio’s many models. Fitting the propensity model on all covariates destabilizes targeting under positivity violations, so it is built collaboratively in three steps (Figure S2): (1) a double-selection screen ^21^ restricts candidates to covariates predictive of both treatment and outcome; (2) survivors are ranked by their approximate contribution to the estimator’s bias term; and (3) nested propensity models over the ordered prefixes of this ranking are compared by cross-validated negative log-likelihood, and the prefix minimizing held-out loss is selected — or an intercept-only model if none improves. Comparisons span two to six arms.

For multi-arm treatments we estimate each arm’s mean and report pairwise contrasts against a common reference. The estimand is a difference in treatment-specific means for continuous and count outcomes and a risk difference for binary outcomes. We report Benjamini–Hochberg FDR q-values (within outcome-by-window families) alongside unadjusted p-values but do not impose family-wise control, impractical and arbitrary at this scale ^22,23^. Significance flags and stratified-report selection use uncorrected p-values to surface even marginal signals, with q-values showing what survives correction. All findings are treated as exploratory, and we additionally computed unadjusted comparisons as a transparency benchmark. Per-arm propensity overlap is examined for every contrast, with positivity diagnostics in each scenario’s evidence report.

Full estimation details are in Supplementary Information S1, Sections 8–9.

#### Effect-Measure Modification and Subgroup Analysis

To generate subgroup evidence we produced stratum-specific estimates for each baseline candidate effect modifier — a median of 185 per scenario (range 176–210) — the 28 Elixhauser comorbidities, baseline laboratory measures and their threshold indicators, adverse-event history, utilization measures, and demographics. Binary modifiers defined two strata.

Continuous modifiers were split at the sample median, giving a prespecified, uniform two-stratum contrast across all candidates (finer effect-modification modeling is future work). Within each stratum we applied the primary Ju C-TMLE (and the unadjusted benchmark). Candidate modifiers were ranked by an effect-measure-modification test — whether the between-arm contrast is homogeneous across strata — using a precision-weighted chi-square on the arm-specific stratum effects ^24^ (degrees of freedom equal to the number of arms minus one; full derivation in Supplementary Information S1, Section 10). A modifier is surfaced as a dedicated subgroup report only when the test is significant (p < 0.05) and at least one stratum-specific contrast exceeds a pre-specified clinical-relevance threshold for some outcome. Effect-modification tests are additionally Benjamini–Hochberg corrected. Because each modifier is screened against hundreds of outcome-by-window tests, the “significant for at least one outcome” criterion remains permissive even after correction, so the number of flagged modifiers reflects screening yield rather than a count of established modifiers. All subgroup findings are treated as exploratory ^25,26^.

### Automation of Narrative Summaries and Quality Control

Quantitative content (estimates, confidence intervals, sample sizes, significance flags) is rendered deterministically from the estimate tables. Large language models only translate encoded outcome and subgroup names into plain language and compose the narrative overview, so numerical transcription is never delegated to the model (Supplementary Information S1, Section 11). Because summarization is only as reliable as the underlying estimates, statistical outputs passed through three-stage quality control before narrative conversion: (1) automated checks for missingness, out-of-range values, and internal inconsistencies; (2) clinical review for face validity; and (3) statistical review for specification fidelity, uncertainty quantification, and appropriate framing of multiplicity. This redundancy is deliberate: conventional peer review cannot catch statistical errors across millions of interconnected estimates ^27–29^. The narrative layer is an engineering component, not a validated model contribution: the reported numbers are guaranteed by deterministic rendering, while formal accuracy evaluation of the generated prose against a manually adjudicated reference set is deferred to companion work.

## Results

Every scenario ran end to end without per-question analyst intervention. We first describe the scale of the output, then benchmark one contrast against a randomized trial.

### Scope and Multi-Stakeholder Coverage

Across 135 clinical scenarios, the workflow generated 210,584,509 outcome evaluations (Figure 3). An evaluation is one outcome × follow-up window × treatment contrast × population stratum × estimator with uncertainty bounds and diagnostics. These are computational outputs, not independent tests or distinct findings, comparable in intent (though not estimand) to prior large-scale efforts ^9,10,30^. Each scenario covered 1,038 distinct outcome measures spanning clinical events, healthcare-resource utilization, and adverse events. The three windows, contrasts, strata, and two estimators multiply this to the portfolio total. The workflow screened a median of 185 baseline features per scenario for effect-measure modification, preserving population-average estimates while surfacing clinically relevant heterogeneity ^23,25,26^. Approximately 7,500 narrative summaries passed through the three-stage quality-control workflow before dissemination ^4,27^.

### Benchmarking a Head-to-Head Contrast Against a Randomized Trial

Our tirzepatide-versus-semaglutide comparison in obese patients can be benchmarked against SURMOUNT-5, the randomized trial of these agents ^31^, which enrolled approximately 750 adults with obesity without diabetes and reported significant superiority of tirzepatide on its primary and five secondary weight endpoints. In our real-world BMI ≥ 35 population (1,318 tirzepatide vs 3,095 high-dose semaglutide initiators) — broader and more representative than SURMOUNT-5’s diabetes-free, BMI ≥ 30 cohort — the adjusted C-TMLE analysis evaluated the same contrast across 155 endpoints and up to three windows (461 endpoint-by-window estimates).

Baseline characteristics for both arms are in the unstratified evidence report (Supplement S2). Continuous-outcome analyses are complete-case, so per-endpoint sample sizes fall below the initiator counts (e.g., n ≈ 1,206 for the 3–12-month BMI contrast). Our analysis agreed with the trial’s central clinical finding: greater adiposity reduction with tirzepatide. The adjusted between-arm BMI difference was −0.85 kg/m² at 3–12 months and −1.02 kg/m² at 12–24 months (both q < 0.05). The real-world magnitude was smaller than in the trial, as expected given the BMI ≥ 35 population, real-world adherence and dosing, and the as-treated estimand. The contrast also spans tirzepatide’s rapid post-2022 uptake, so it is subject to secular-trend confounding. Consistent with the trial, no significant difference appeared in serious safety events (major adverse cardiac events, pancreatitis, gallbladder disease, chronic kidney disease; all q > 0.20).

This concordance warrants caution, because these adverse-event contrasts reflect on-treatment observation rather than incidence (see Limitations). Across the full panel, 34 of 461 estimates (7.4%) were significant after Benjamini–Hochberg control, spanning 27 endpoints beyond the trial’s six — glycemic, lipid, renal, hepatic, and hematologic trajectories, gastrointestinal tolerability, utilization, and treatment persistence. Tirzepatide also showed a lower incidence of stage 2 hypertension (diastolic blood pressure ≥ 90 mmHg; risk difference −0.04, q < 0.001) and a modest reduction in systolic blood pressure (−1.3 mmHg, q = 0.05) versus semaglutide. The paired effect-modification screen flagged 60 baseline features as significant modifiers of at least one treatment–outcome relationship, each a hypothesis-generating subgroup analysis.

Tirzepatide’s advantage on the primary weight endpoint proved robust to stratification: across 1,312 adequately-powered subgroup estimates spanning 178 baseline effect modifiers, it remained directionally superior in all but one and was not significantly reversed in any — subgroups the trial was not powered to examine. On secondary endpoints, by contrast, the screen surfaced discordant subgroups: although tirzepatide reduced the rate of morbid obesity (BMI ≥ 40) overall at 3–12 months, this benefit reversed among patients with baseline insomnia (adjusted risk difference +0.06 [95% CI +0.01, +0.12]; effect-modification q < 0.001), a signal of heterogeneous response.

## Discussion

This paper describes a framework for high-throughput observational evidence from linked EHR and claims data: clinician-selected scenarios, scalable cohort construction, and a pre-specified measurement architecture covering 1,038 outcomes per scenario. The central contribution is not any individual finding but a different way of producing evidence that reduces fragmentation by standardizing what is measured, across how many outcomes, and for which stakeholders, before results are known. Because the architecture is fixed, adding a scenario reuses the same measurement, estimation, and quality-control machinery, so the marginal cost of new evidence is low.

Fragmentation in observational research is primarily a measurement problem, not a statistical one. Prior literature is contradictory not because groups used different estimators but because they measured different outcomes over different windows in differently defined populations. Pre-specifying a common set of outcomes applied identically across all 135 scenarios makes cross-domain comparison and shared, multi-stakeholder interpretation possible.

Standardization also makes visible what single studies cannot: divergence between adjusted and unadjusted estimates flags where confounding is most consequential, and population-average effects hide clinically meaningful variation. Our workflow screens baseline features for differential response and generates subgroup estimates for populations trials routinely exclude, moving the question from “does A beat B?” to “for which patients, under which circumstances, and by how much?” — what precision medicine requires.

Our approach shares intent with prior large-scale efforts reducing selective reporting through prespecified execution across many hypotheses ^9–12^ and with collaborative network studies built on common data models ^32–34^. We differ in using linked EHR–claims data integrated into a single dataset, a doubly robust collaborative TMLE as the primary estimator with an unadjusted benchmark, and multi-stakeholder outcome panels with structured narrative summaries.

Multiplicity is unavoidable at this scale but secondary: residual confounding and measurement error dominate bias in observational data ^6,35^. We treat all findings as exploratory, interpreting them in aggregate rather than as isolated tests.

Several limitations warrant emphasis. Continuous-outcome analyses are complete-case, valid only under missingness-at-random given measured covariates ^36^. Labs are ordered in response to symptoms, so patients with an observed outcome are not a random subset of the cohort. If that selection differs by arm, complete-case estimates are biased. We do not yet reweight by the probability of being measured. Censoring is a further limitation. Follow-up ends at treatment discontinuation or switching, and both are driven by tolerability and response, so censoring is arm-dependent. Treating this as a point-treatment problem is therefore biased: censoring acts as a time-varying intervention alongside treatment, and baseline adjustment cannot remove it. IPCW-TMLE and related time-varying methods are the planned remedy in the companion validation. For binary outcomes, unrecorded events are treated as non-events, conflating true absence with unobserved follow-up. This is most consequential for adverse events, where worse-tolerated arms accrue less on-treatment observation, so adverse-event risk differences are not incidence contrasts. Phenotypes defined by code lists are not yet formally validated; secular trends may confound comparisons involving newer therapies; and downsampling to 5,000 per arm trades precision and can shift the effective population. Reported q-values do not yet gate which findings are surfaced. External groups cannot reproduce these estimates from the source data (see Data availability). This manuscript reports the workflow rather than a formal validation. Systematic calibration against negative and positive controls ^37^ and coverage under realistic conditions are ongoing companion work.

The tirzepatide–semaglutide example shows how generated evidence complements rather than competes with trials: our analysis was consistent with SURMOUNT-5’s result in a BMI ≥ 35 population trials exclude, while characterizing more than 150 additional endpoints the trial was not built to address.

In sum, the value of this work lies in its measurement architecture: one pre-specified design of outcomes, baselines, and windows serves many stakeholders. Coupling comparative effectiveness with systematic effect-modification screening begins to move observational evidence from what works on average toward what works for whom — an exploratory step toward precision medicine at the scale of routine care.

## Acknowledgements

Alan Hubbard was paid as a consultant for his contributions to this project and publication.

Use of large language models. Large language models (Claude Opus 5 and Claude Sonnet 5, Anthropic, accessed via the Claude Code interface) were used to assist in drafting and editing the text of this manuscript, including composing and revising prose from author-supplied outlines, results, and source material, and copyediting for clarity and concision. The models were not used to generate, analyze, or transcribe any study data, statistical estimates, or figures; all quantitative content derives from the analytic pipeline described in Methods. All authors carefully reviewed, verified, and approved every passage of AI-assisted text and take full responsibility for the accuracy, integrity, and originality of the submitted work.

## Data availability

Our data-use agreements prohibit redistribution of the underlying data, so external groups cannot reproduce these specific estimates. Complete measurement, cohort, and analytic specifications are provided in Supplementary Information S1 (Sections 1–10) to enable replication on other mapped datasets.

---

Figure 2. Example Summary From the Bariatric Surgery and Tirzepatide vs Semaglutide Case (unstratified).

**While the generated reports can be hundreds of pages, we provide these short, easy-to-digest, summaries at the start, which makes these more consumable to a clinical audience. Persistence and discontinuation-based measures apply to ongoing pharmacotherapy and are not meaningful for one-time procedures such as bariatric surgery. The adjusted estimator is the collaborative TMLE (C-TMLE) described in Methods.**

We conducted a high-throughput observational study using linked electronic health record and claims data to estimate the effects of bariatric surgery and tirzepatide high dose compared to semaglutide high dose on a broad set of outcomes across heterogeneous patient populations. The analysis evaluated multiple outcome domains, including clinical effectiveness, adverse events, and healthcare utilization and costs, while characterizing variation in effects across patient demographics, comorbid conditions, prior procedures, and medication exposures. For each outcome, we applied targeted maximum likelihood estimation (TMLE) to adjust for confounding and minimize model misspecification. This approach enabled the simultaneous generation of robust, population-level and subgroup-specific estimates across a large number of clinically relevant endpoints, providing a comprehensive view of treatment benefits, risks, and economic impacts in real-world clinical practice. Mean Difference (MD) is used for continuous outcomes, Risk Difference (RD) for binary outcomes, and Effect Measure Modification (EMM) describes differences in treatment effects across subgroups; for all metrics, a negative value indicates a lower incidence or value in the treatment arm compared to the reference arm. Compared with semaglutide high dose, bariatric surgery led to a lower body-mass index (BMI) (MD −2.80 [−2.93, −2.67], n=2,080) and a lower change in BMI (MD −2.99 [−3.21, −2.76], n=2,077) at 0-3 months, with these reductions sustained at 3-12 months for both BMI (MD −5.94 [−6.13, - 5.76], n=2,887) and change in BMI (MD −5.91 [−6.16, −5.66], n=2,883). Similarly, compared with semaglutide high dose, tirzepatide high dose led to a lower BMI (MD −0.85 [−1.11, −0.59], n=1,206) and a lower change in BMI (MD −1.05 [−1.49, −0.61], n=1,206) at 3-12 months, as well as a lower change in BMI at 0-3 months (MD −0.66 [−1.03, −0.30], n=879). There were no significant differences between either treatment arm and semaglutide high dose regarding the counts of cholelithiasis and cholecystitis, chronic kidney disease, major adverse cardiac events, or pancreatitis at 0-3 or 3-12 months. Compared with semaglutide high dose, bariatric surgery and tirzepatide high dose led to a lower body weight at 3-12 months (MD −36.00 [−38.70, - 33.30], n=2,749; MD −4.50 [−8.23, −0.77], n=1,114, respectively), a lower change in body weight (MD −35.10 [−37.00, −33.10], n=2,688; MD −5.24 [−7.94, −2.55], n=1,095, respectively), and a lower rate of obesity (BMI > 30) (RD −0.17 [−0.19, −0.15], n=3,095; RD −0.04 [−0.07, −0.01], n=1,318, respectively). Finally, regarding treatment persistence at 3-12 months, patients receiving bariatric surgery remained on therapy for an absolute duration of 263.0 days (n=3,095). Note: All intervals presented in brackets represent 95% Confidence Intervals.

Figure 3. Analysis-Count Cascade. The portfolio’s 210,584,509 outcome evaluations arise from 135 clinician-selected scenarios × approximately 1,038 outcome measures per scenario × 3 post-index windows (0–90, 91–365, 366–730 days) × treatment contrasts (2–6 arms) × 2 estimators (adjusted collaborative TMLE and an unadjusted benchmark), together with a subgroup layer screening a median of ≈185 baseline effect modifiers per scenario. Counts vary by scenario; multipliers are approximate.

## References

1. He J, Morales DR, Guthrie B. Exclusion rates in randomized controlled trials of treatments for physical conditions: a systematic review. Trials 2020;21(1):228.

2. Corrigan-Curay J, Sacks L, Woodcock J. Real-World Evidence and Real-World Data for Evaluating Drug Safety and Effectiveness. JAMA 2018;320(9):867–8.

3. Franklin JM, Liaw K-L, Iyasu S, Critchlow CW, Dreyer NA. Real-world evidence to support regulatory decision making: New or expanded medical product indications. Pharmacoepidemiology and Drug Safety 2021;30(6):685–93.

4. Ostropolets A, Albogami Y, Conover M, others. Reproducible variability: assessing investigator discordance across 9 research teams attempting to reproduce the same observational study. Journal of the American Medical Informatics Association 2023;30(5):859– 68.

5. Leducq S, Zaki F, Hollestein LM, others. The majority of observational studies in leading peer-reviewed medicine journals are not registered and do not have a publicly accessible protocol: a scoping review. Journal of Clinical Epidemiology 2024;170:111341.

6. Schuemie MJ, Ryan PB, DuMouchel W, Suchard MA, Madigan D. Interpreting observational studies: why empirical calibration is needed to correct p-values. Statistics in Medicine 2014;33(2):209–18.

7. Dang LE, Gruber S, Lee H, others. A Causal Roadmap for Generating High-Quality Real-World Evidence. Journal of Clinical Epidemiology 2024;

8. Petersen ML, van der Laan MJ. Causal models and learning from data: integrating causal modeling and statistical estimation. Epidemiology 2014;25(3):418–26.

9. Schuemie MJ, Ryan PB, Pratt N, others. Principles of Large-scale Evidence Generation and Evaluation across a Network of Databases (LEGEND). Journal of the American Medical Informatics Association 2020;27(8):1331–7.

10. Schuemie MJ, Ryan PB, Hripcsak G, Madigan D, Suchard MA. Improving reproducibility by using high-throughput observational studies with empirical calibration. Philosophical Transactions of the Royal Society A 2018;376(2128):20170356.

11. Platt R, Brown JS, Robb M, others. The FDA Sentinel Initiative — An Evolving National Resource. New England Journal of Medicine 2018;379(22):2091–3.

12. Tatonetti NP, Ye PP, Daneshjou R, Altman RB. Data-driven prediction of drug effects and interactions. Science Translational Medicine 2012;4(125):125ra31.

13. Benchimol EI, Smeeth L, Guttmann A, others. The REporting of studies Conducted using Observational Routinely-collected health Data (RECORD) statement. PLoS Medicine 2015;12(10):e1001885.

14. von Elm E, Altman DG, Egger M, Pocock SJ, Götzsche PC, Vandenbroucke JP. The Strengthening the Reporting of Observational Studies in Epidemiology (STROBE) statement: guidelines for reporting observational studies. The Lancet 2007;370(9596):1453–7.

15. Hernán MA, Robins JM. Using Big Data to Emulate a Target Trial When a Randomized Trial Is Not Available. American Journal of Epidemiology 2016;183(8):758–64.

16. Elixhauser A, Steiner C, Harris DR, Coffey RM. Comorbidity measures for use with administrative data. Medical Care 1998;36(1):8–27.

17. Quan H, Sundararajan V, Halfon P, others. Coding algorithms for defining comorbidities in ICD-9-CM and ICD-10 administrative data. Medical Care 2005;43(11):1130–9.

18. Li Z, Gogia S, Tatem KS, et al. Developing a Model to Predict High Health Care Utilization Among Patients in a New York City Safety Net System. Medical Care 2023;61(2):102–8.

19. Figueroa JF, Zhou X, Jha AK. Characteristics And Spending Patterns Of Persistently High-Cost Medicare Patients. Health Affairs 2019;38(1):107–14.

20. Ju C, Gruber S, Lendle SD, others. Scalable collaborative targeted learning for high-dimensional data. Statistical Methods in Medical Research 2019;28(2):532–54.

21. Belloni A, Chernozhukov V, Hansen C. Inference on treatment effects after selection among high-dimensional controls. Review of Economic Studies 2014;81(2):608–50.

22. Perneger TV. What’s wrong with Bonferroni adjustments. BMJ 1998;316(7139):1236–8.

23. Glickman ME, Rao SR, Schultz MR. False discovery rate control is a recommended alternative to Bonferroni-type adjustments in health studies. Journal of Clinical Epidemiology 2014;67(8):850–7.

24. VanderWeele TJ, Knol MJ. A Tutorial on Interaction. Epidemiologic Methods 2014;3(1):33–72.

25. Burke JF, Sussman JB, Kent DM, Hayward RA. Three simple rules to ensure reasonably credible subgroup analyses. BMJ 2015;351:h5651.

26. Wang R, Lagakos SW, Ware JH, Hunter DJ, Drazen JM. Statistics in medicine— reporting of subgroup analyses in clinical trials. New England Journal of Medicine 2007;357(21):2189–94.

27. Tennant JP, Ross-Hellauer T. The limitations to our understanding of peer review. Research Integrity and Peer Review 2020;5:6.

28. Bacchetti P. Peer review of statistics in medical research: the other problem. BMJ 2002;324(7348):1271–3.

29. Strasak AM, Zaman Q, Pfeiffer KP, Göbel G, Ulmer H. Statistical errors in medical research - a review of common pitfalls. Swiss Medical Weekly 2007;137(3–4):44–9.

30. Suchard MA, Schuemie MJ, Krumholz HM, others. Comprehensive comparative effectiveness and safety of first-line antihypertensive drug classes: a systematic, multinational, large-scale analysis. The Lancet 2019;394(10211):1816–26.

31. Aronne LJ, others. Tirzepatide versus Semaglutide for the Treatment of Obesity (SURMOUNT-5). New England Journal of Medicine 2025;

32. Hripcsak G, Duke JD, Shah NH, others. Observational Health Data Sciences and Informatics (OHDSI): Opportunities for Observational Researchers. Studies in Health Technology and Informatics 2015;216:574–8.

33. Hripcsak G, Ryan PB, Duke JD, others. Characterizing treatment pathways at scale using the OHDSI network. Proceedings of the National Academy of Sciences 2016;113(27):7329–36.

34. Overhage JM, Ryan PB, Reich CG, Hartzema AG, Stang PE. Validation of a common data model for active safety surveillance research. Journal of the American Medical Informatics Association 2012;19(1):54–60.

35. Funk MJ, Landi SN. Misclassification in administrative claims data: quantifying the impact on treatment effect estimates. Current Epidemiology Reports 2014;1(4):175–85.

36. Little RJA, Rubin DB. Statistical Analysis with Missing Data. 3rd ed. Hoboken, NJ: Wiley; 2019.

37. Observational Health Data Sciences and Informatics (OHDSI). Method Validity [Internet]. In: The Book of OHDSI. 2021. Available from: https://ohdsi.github.io/TheBookOfOhdsi/MethodValidity.html

