## Supplemental S1 for "High-Throughput Observational Evidence Generation Using Linked Electronic Health Record and Claims Data"

**Methods: Measurement Framework for Real-World Evidence Comparative Effectiveness Studies**

This supplement provides the complete measurement and analytic specifications for the main manuscript, detailing the observation windows, baseline covariates, and outcome definitions (Sections 1–7) and the unadjusted sensitivity analysis (Section 8) referenced there.

### Part A — Measurement Architecture

#### 1. Study Design and Observation Windows

This retrospective cohort study utilized real-world data (RWD) from electronic health records (EHR) and administrative claims. The study design employs an active-comparator, new-user framework. The index date is defined as the date of the first qualifying prescription fill for the study medication or, for procedure- or device-based comparisons, the first qualifying procedure or service date. All time windows are anchored to this index date. Patients were required to have a minimum of 3 months of enrollment history prior to the index date and a minimum of 6 months of post-index follow-up. Follow-up was censored at the earliest of death, disenrollment, discontinuation of the index treatment, or switch to a second-line agent, so outcomes were ascertained within an on-treatment (as-treated) window.

##### **1.1 Baseline Assessment Period**

Baseline characteristics were assessed in a single pre-index window. All laboratory values, vital signs, comorbidity burden, healthcare utilization patterns, and adverse event history were measured during this period to establish pre-treatment status.

| **Window Label** | **Start** | **End** | **Description** |
| --- | --- | --- | --- |
| 365-to-15 days pre-index | Index date minus 1 year | Index date minus 15 days | Primary baseline window; the 15-day washout before index avoids capturing peri-prescribing lab draws that may reflect early treatment effects |

##### **1.2 Outcome Assessment Periods**

Post-index outcomes were assessed across three windows (Figure S1) to capture acute, intermediate, and long-term treatment effects. The 0–90 day window serves as a cumulative early-phase summary, while the 91–365 and 366–730 day windows enable period-specific comparisons.

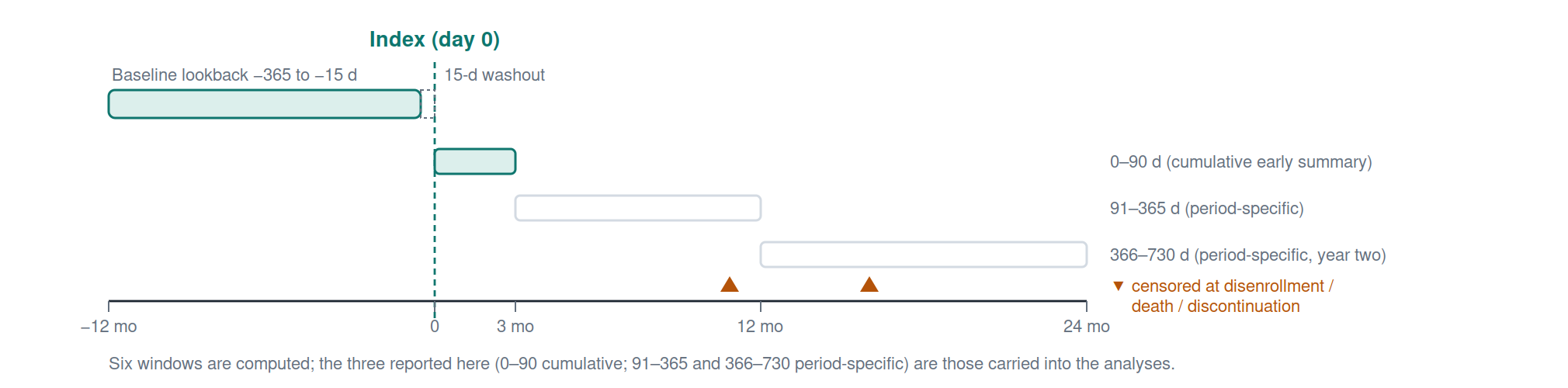

Figure S1. Cohort observation-window timeline. Schematic of the index date (day 0), the 365-to-15-day baseline lookback with a 15-day washout, and the three reported post-index outcome windows — 0–90 days (cumulative early summary) and 91–365 and 366–730 days (period-specific) — with follow-up censored at the earliest of disenrollment, death, or treatment discontinuation. Six windows are computed; the three shown are those carried into the analyses.

| **Window Label** | **Start** | **End** | **Purpose** |
| --- | --- | --- | --- |
| Day 0–90 | Index date | Index date + 90 days | Cumulative early-phase summary (acute + short-term) |
| Day 91–365 | Index date + 91 days | Index date + 365 days | Intermediate-term efficacy, durability, and safety |
| Day 366–730 | Index date + 366 days | Index date + 730 days | Long-term outcomes in year two of therapy |

##### **1.3 Change-from-Baseline (Delta) Calculations**

For continuous laboratory and vital sign measures, change-from-baseline (Δ) values were computed as the difference between the last observed value in a given outcome window and the last observed value in the 365-to-15-day baseline window. Deltas were calculated for three outcome windows: 0–90 days, 91–365 days, and 366–730 days. Clinically plausible delta ranges were enforced for each measure to exclude erroneous values.

#### 2. Baseline Comorbidity Assessment

Comorbidity burden was assessed at baseline using the 28 individual Elixhauser comorbidity indicators (separate binary flags, not a weighted summary index), which identify 28 distinct clinical conditions from International Classification of Diseases, Tenth Revision (ICD-10) and International Classification of Diseases, Ninth Revision (ICD-9) diagnosis codes. Each comorbidity was evaluated as a binary indicator (present/absent) during the 365-to-15-day pre-index baseline window. The following conditions were assessed:

| **Condition** | **Condition** | **Condition** | **Condition** |
| --- | --- | --- | --- |
| Acquired Immunodeficiency Syndrome (AIDS) / Human Immunodeficiency Virus (HIV) | Alcohol abuse | Anemia (deficiency) | Blood loss anemia |
| Cardiac arrhythmia | Coagulopathy | Congestive heart failure (CHF) | Depression |
| Diabetes mellitus, complicated | Diabetes mellitus, uncomplicated | Drug abuse | Fluid and electrolyte disorders |
| Hypertension, complicated | Hypertension, uncomplicated | Hypothyroidism | Liver disease |
| Lymphoma | Metastatic cancer | Obesity | Other neurological disorders |
| Paralysis | Peptic ulcer disease | Peripheral vascular disorders | Psychoses |
| Rheumatoid arthritis / collagen vascular diseases | Solid tumor without metastasis | Valvular disease | Weight loss |

#### 3. Healthcare Utilization

Healthcare resource use was quantified across both the baseline and all outcome windows. Each utilization category was measured as a binary indicator (any use vs. no use) as well as a continuous measure: visit count for encounter-based categories or length of stay (LOS) in days for facility-based categories. The following 14 utilization categories were assessed:

| **Utilization Category** | **Identification Method** | **Continuous Measure** |
| --- | --- | --- |
| Intensive Care Unit (ICU) | Current Procedural Terminology (CPT) codes: 99291, 99292, 31500, 94002–94004 | Length of stay (days) |
| Inpatient hospitalization | Visit type flags (Inpatient Visit, Inpatient Hospital Stay, ER-to-Inpatient) and CPT evaluation & management (E/M) codes: 99221–99239 | Length of stay (days) |
| Skilled nursing facility (SNF) | CPT codes: 99304–99310, 99315–99316 | Visit count |
| Home / domiciliary visits | CPT codes: 99341–99350 | Visit count |
| Urgent care (combined with ED) | CPT billing codes: S9083, S9088 (combined with ED visits) | Visit count |
| Emergency department (ED) | CPT E/M codes: 99281–99285 and Emergency Room visit type flags | Visit count |
| Outpatient / office visits | CPT E/M codes: 99202–99205, 99212–99215 and Office Visit type flags | Visit count |
| Common diagnostic imaging | CPT codes for mammography, chest X-ray, dual-energy X-ray absorptiometry (DEXA), magnetic resonance imaging (MRI), ultrasound, computed tomography (CT) | Visit count |
| Basic laboratory workup | CPT codes: 80048 (basic metabolic panel [BMP]), 80053 (comprehensive metabolic panel [CMP]), 80061 (lipid panel), 85025/85027 (complete blood count [CBC]), 84443 (thyroid-stimulating hormone [TSH]), 83036 (hemoglobin A1c [HbA1c]) | Visit count |
| Physical therapy (PT) / Occupational therapy (OT) | CPT codes: 97010–97799 (therapeutic procedures, evaluations, functional assessments) | Visit count |
| Psychotherapy | CPT codes: 90832–90853 (individual, family, and group psychotherapy) | Visit count |
| Preventive / wellness visits | CPT codes: 99381–99397 (preventive E/M, new and established patients) | Visit count |
| Transitional care management (TCM) | CPT codes: 99495–99496 | Visit count |
| Advance care planning (ACP) | CPT codes: 99497–99498 | Visit count |

#### 4. Laboratory and Vital Sign Measurements

Continuous laboratory and vital sign values were extracted using Logical Observation Identifiers Names and Codes (LOINC) identifiers from structured EHR data. Each measure was filtered to a clinically plausible range to exclude implausible or erroneous values. For each time window, the last observed value (LAST MENTION) was used as the representative measurement. The following 30 measures were captured:

##### **4.1 Hepatic Function**

| **Measure** | **LOINC Code(s)** | **Plausible Range** | **Unit** |
| --- | --- | --- | --- |
| Alanine aminotransferase (ALT/SGPT) | 1742-6, 1743-4, 1744-2 | 2–10,000 | U/L |
| Aspartate aminotransferase (AST/SGOT) | 1920-8, 30239-8, 88112-8 | 2–10,000 | U/L |
| Gamma-glutamyl transferase (GGT) | 2324-2 | 3–5,000 | U/L |
| Albumin | 1751-7, 61151-7, 61152-5, 101198-0 | 0.5–7.0 | g/dL |
| Total bilirubin | 1975-2 | 0.1–50 | mg/dL |
| Total protein | 2885-2 | 2–14 | g/dL |

##### **4.2 Renal Function**

| **Measure** | **LOINC Code(s)** | **Plausible Range** | **Unit** |
| --- | --- | --- | --- |
| Creatinine | 38483-4, 2160-0 | 0.1–20 | mg/dL |
| Estimated glomerular filtration rate (eGFR) | 33914-3, 48642-3, 48643-1, 50210-4, 62238-1, 69405-9, 98979-8, 76633-7, 77147-7, 98980-6, 88293-6, 88294-4 | 5–180 | mL/min/1.73m² |
| Blood urea nitrogen (BUN) | 3094-0, 6299-2 | 1–200 | mg/dL |
| Albumin-to-creatinine ratio (ACR) | 9318-7 | 1–10,000 | mg/g |

##### **4.3 Lipid Panel**

| **Measure** | **LOINC Code(s)** | **Plausible Range** | **Unit** |
| --- | --- | --- | --- |
| Total cholesterol | 2093-3 | 60–700 | mg/dL |
| Low-density lipoprotein (LDL) | 2089-1, 13457-7, 18262-6, 55440-2, 18261-8 | 20–400 | mg/dL |
| High-density lipoprotein (HDL) | 2085-9 | 10–150 | mg/dL |
| Triglycerides (TG) | 2571-8 | 10–5,000 | mg/dL |

##### **4.4 Glycemic / Metabolic**

| **Measure** | **LOINC Code(s)** | **Plausible Range** | **Unit** |
| --- | --- | --- | --- |
| Hemoglobin A1c (HbA1c) | 4548-4, 4549-2, 17856-6 | 3.5–18 | % |
| Blood glucose | 2345-7, 74774-1, 2339-0, 2341-6, 6777-7 | 25–1,000 | mg/dL |

##### **4.5 Hematologic**

| **Measure** | **LOINC Code(s)** | **Plausible Range** | **Unit** |
| --- | --- | --- | --- |
| Hemoglobin | 718-7, 20509-6, 30313-1, 14775-1, 30350-3, 76769-9, 30352-9 | 2–23 | g/dL |
| Platelets | 777-3, 26515-7 | 1–1,000 | K/µL |
| White blood cells (WBC) / leukocytes | 20584-9, 6690-2, 26464-8 | 0.5–500 | K/µL |
| Prothrombin time – International Normalized Ratio (PT-INR) | 34714-6, 6301-6, 46418-0, 5895-7, 38875-1 | 0.1–20 | INR |

##### **4.6 Inflammation / Acute Phase Reactants**

| **Measure** | **LOINC Code(s)** | **Plausible Range** | **Unit** |
| --- | --- | --- | --- |
| High-sensitivity C-reactive protein (hs-CRP) / CRP | 30522-7, 71426-1, 76486-0 | 0.1–500 | mg/L |
| C-reactive protein (CRP), standard | 1988-5 | 0.1–500 | mg/L |

##### **4.7 Cardiovascular / Cardiac Biomarkers**

| **Measure** | **LOINC Code(s)** | **Plausible Range** | **Unit** |
| --- | --- | --- | --- |
| N-terminal pro-B-type natriuretic peptide (NT-proBNP) | 33762-6, 83107-3 | 5–50,000 | pg/mL |

##### **4.8 Thyroid Function**

| **Measure** | **LOINC Code(s)** | **Plausible Range** | **Unit** |
| --- | --- | --- | --- |
| Thyroid-stimulating hormone (TSH) | 3016-3, 11580-8, 11579-0 | 0.01–100 | mIU/L |

##### **4.9 Micronutrients and Electrolytes**

| **Measure** | **LOINC Code(s)** | **Plausible Range** | **Unit** |
| --- | --- | --- | --- |
| Vitamin D, serum (25-hydroxyvitamin D) | 35365-6, 62292-8 | 1–200 | ng/mL |
| Potassium | 2823-3 | 0.6–15 | mEq/L |

##### **4.10 Anthropometrics and Vital Signs**

| **Measure** | **LOINC Code(s)** | **Plausible Range** | **Unit** |
| --- | --- | --- | --- |
| Body mass index (BMI) | 39156-5, 41909-3, 89270-3 | 13–80 | kg/m² |
| Body weight | 29463-7 | 1–600 | lbs |
| Systolic blood pressure (SBP) | 8480-6 | 50–280 | mmHg |
| Diastolic blood pressure (DBP) | 8462-4 | 30–160 | mmHg |

#### 5. Clinically Meaningful Laboratory and Vital Sign Thresholds (Binary)

In addition to continuous values, each laboratory and vital sign measure was dichotomized at one or more clinically meaningful thresholds. These binary indicators capture the proportion of patients exceeding or falling below guideline-endorsed or clinically significant cutpoints during each time window. The threshold value and directionality are embedded in each variable definition.

##### **5.1 Hepatic Function Thresholds**

| **Threshold** | **Definition** | **Cutpoint** |
| --- | --- | --- |
| Elevated ALT | ALT > 3x upper limit of normal (ULN) | >120 U/L |
| Elevated AST | AST > 3x ULN | >120 U/L |
| Elevated GGT | GGT above normal range | >100 U/L |
| Transaminitis (composite) | Elevated ALT or elevated AST (union) | Either >120 U/L |
| Reduced albumin (hypoalbuminemia) | Albumin below normal | <3.5 g/dL |
| Elevated total bilirubin (hyperbilirubinemia) | Bilirubin above normal | >1.3 mg/dL |
| Low total protein (hypoproteinemia) | Total protein below normal | <6.0 g/dL |
| Elevated total protein (hyperproteinemia) | Total protein above normal | >8.5 g/dL |

##### **5.2 Renal Function Thresholds**

| **Threshold** | **Definition** | **Cutpoint** |
| --- | --- | --- |
| Elevated creatinine | Creatinine above normal | >1.3 mg/dL |
| Reduced eGFR | Chronic Kidney Disease (CKD) stage 3+ threshold | <60 mL/min/1.73m² |
| Elevated ACR (proteinuria) | Macroalbuminuria | >300 mg/g |
| Elevated BUN (mild) | BUN mildly elevated | >25 mg/dL |
| Elevated BUN (severe) | BUN severely elevated | >50 mg/dL |

##### **5.3 Lipid Panel Thresholds**

| **Threshold** | **Definition** | **Cutpoint** |
| --- | --- | --- |
| Elevated total cholesterol | Hypercholesterolemia | >240 mg/dL |
| LDL at very high-risk goal | Per ACC/AHA guidelines for very high-risk patients | <69 mg/dL |
| LDL at high-risk goal | Per ACC/AHA guidelines for high-risk patients | 70–99 mg/dL |
| LDL at general population goal | Borderline level for general population | 100–129 mg/dL |
| LDL high risk | Elevated LDL | >160 mg/dL |
| LDL very high risk | Severely elevated LDL | >190 mg/dL |
| Protective HDL | HDL above cardioprotective threshold | >60 mg/dL |
| Elevated triglycerides (hypertriglyceridemia) | Triglycerides above normal | >200 mg/dL |

##### **5.4 Glycemic / Metabolic Thresholds**

| **Threshold** | **Definition** | **Cutpoint** |
| --- | --- | --- |
| HbA1c normal | Non-diabetic range | <5.6% |
| HbA1c prediabetes | Prediabetes range per American Diabetes Association (ADA) | 5.7–6.4% |
| HbA1c at goal | Typical glycemic target for diabetic patients | 6.5–7.5% |
| HbA1c elevated | Above target for most patients | >7.6% |
| HbA1c Centers for Medicare & Medicaid Services (CMS) quality flag | CMS quality measure threshold for poor glycemic control | >9.0% |
| Elevated blood glucose (diabetes threshold) | Random glucose exceeding diabetes diagnostic cutpoint | >200 mg/dL |
| Low blood glucose (hypoglycemia) | Clinically significant hypoglycemia | <70 mg/dL |

##### **5.5 Hematologic Thresholds**

| **Threshold** | **Definition** | **Cutpoint** |
| --- | --- | --- |
| Reduced hemoglobin (severe anemia) | Hemoglobin indicating severe anemia | <7 g/dL |
| Low platelets (thrombocytopenia) | Platelets below clinically significant threshold | <50 K/µL |
| Elevated WBC (leukocytosis) | WBC above normal range | >15 K/µL |
| Reduced WBC (leukopenia) | WBC below normal range | <3.0 K/µL |
| Elevated PT-INR (supratherapeutic) | INR above therapeutic range | >3.0 INR |
| Elevated PT-INR (critical) | INR at critical value | >5.0 INR |
| Hypokalemia | Potassium below normal (LOINC 2823-3) | <3.4 mEq/L |
| Hyperkalemia | Potassium above normal (LOINC 2823-3) | >5.1 mEq/L |

##### **5.6 Inflammation / Acute Phase Reactant Thresholds**

| **Threshold** | **Definition** | **Cutpoint** |
| --- | --- | --- |
| hs-CRP low cardiovascular (CV) risk | Below average CV risk | <0.9 mg/L |
| hs-CRP average CV risk | Average CV risk range | 1.0–3.0 mg/L |
| hs-CRP high CV risk | Above average CV risk | >3.1 mg/L |
| Elevated CRP (standard) | CRP indicating significant systemic inflammation | >10 mg/L |

##### **5.7 Cardiovascular, Thyroid, Micronutrient, and Anthropometric Thresholds**

| **Threshold** | **Definition** | **Cutpoint** |
| --- | --- | --- |
| Elevated NT-proBNP | Elevated natriuretic peptide suggesting heart failure | >450 pg/mL |
| TSH – hyperthyroid range | Suppressed TSH | <0.3 mIU/L |
| TSH – hypothyroid range | Elevated TSH | >4.7 mIU/L |
| Vitamin D severe deficiency | 25(OH)D severely deficient | <12 ng/mL |
| Obesity (class I+) | BMI at or above obesity threshold | >30 kg/m² |
| Class II obesity | BMI in class II obesity range | >35 kg/m² |
| Morbid obesity (class III) | BMI at or above class III threshold | >40 kg/m² |
| Underweight | BMI below normal | <18.5 kg/m² |
| SBP hypotension | Systolic blood pressure below normal | <89 mmHg |
| SBP stage 1 hypertension (HTN) | Per ACC/AHA 2017 guidelines | 130–139 mmHg |
| SBP stage 2 HTN | Per ACC/AHA 2017 guidelines | ≥140 mmHg |
| SBP hypertensive crisis | Hypertensive urgency/emergency | >180 mmHg |
| DBP hypotension | Diastolic blood pressure below normal | <59 mmHg |
| DBP stage 1 HTN | Per ACC/AHA 2017 guidelines | 80–89 mmHg |
| DBP stage 2 HTN | Per ACC/AHA 2017 guidelines | ≥90 mmHg |
| DBP hypertensive crisis | Hypertensive urgency/emergency | >120 mmHg |
| Hypotension (composite) | SBP <89 or DBP <59 (union) | SBP <89 or DBP <59 |

#### 6. Side Effects and Adverse Events

Adverse events were identified using ICD-10 diagnosis codes captured in claims and EHR encounter data. Each adverse event was assessed as a binary indicator (present/absent) across all baseline and outcome time windows. A total of 43 adverse event categories were monitored, organized by organ system:

##### **6.1 Gastrointestinal (GI) Adverse Events**

| **Adverse Event** | **ICD-10 Code(s)** | **Description** |
| --- | --- | --- |
| Abdominal pain | R10 | Non-specific abdominal pain |
| Constipation | K58.1, K59.0 | Irritable bowel syndrome (IBS) with constipation and functional constipation |
| Diarrhea | K58.0, K58.2, K59.1, R19.7 | IBS with diarrhea, mixed IBS, functional diarrhea, and diarrhea not otherwise specified (NOS) |
| Nausea and vomiting | R11 | Nausea, vomiting, and unspecified emesis |
| Dyspepsia | K30 | Functional dyspepsia / indigestion |
| Flatulence and bloating | R14 | Abdominal distension and excess gas |

##### **6.2 Cardiovascular Adverse Events**

| **Adverse Event** | **ICD-10 Code(s)** | **Description** |
| --- | --- | --- |
| Palpitations | R00.2 | Subjective sensation of abnormal heartbeat |
| Tachycardia | R00.0 | Elevated heart rate |
| Bradycardia | R00.1 | Reduced heart rate |
| Peripheral edema | R60.0 | Localized edema of extremities |
| Flushing | R23.2 | Skin flushing / erythema |
| Hypotension (diagnosis) | I95 | Symptomatic low blood pressure |
| Hypokalemia (diagnosis) | E87.6 | Low serum potassium (diagnostic code) |
| Hyperkalemia (diagnosis) | E87.5 | Elevated serum potassium (diagnostic code) |

##### **6.3 Neurological and Psychiatric Adverse Events**

| **Adverse Event** | **ICD-10 Code(s)** | **Description** |
| --- | --- | --- |
| Dizziness | R42 | Dizziness and giddiness |
| Headaches | G43, G44, R51 | Migraine, tension-type headache, and headache NOS |
| Insomnia | F51.0, G47.0, Z73.81 | Sleep-onset and maintenance insomnia |
| Drowsiness / somnolence | R40.0 | Excessive daytime sleepiness |
| Fatigue | R53.83 | Generalized fatigue and malaise |
| Tremor | R25.1, G25.0 | Essential and non-specific tremor |
| Peripheral neuropathy | G62.9 | Unspecified polyneuropathy |
| Anxiety | F41, F41.1, F41.9 | Generalized and unspecified anxiety disorders |
| Depression | F32, F33 | Major depressive disorder (MDD), single and recurrent episodes |
| Confusion / cognitive changes | R41.0, R41.3 | Disorientation, altered cognition |

##### **6.4 Metabolic / Electrolyte Adverse Events**

| **Adverse Event** | **ICD-10 Code(s)** | **Description** |
| --- | --- | --- |
| Hyponatremia | E87.1 | Low serum sodium |
| Hypoglycemia | E16.0, E16.2 | Drug-induced and other hypoglycemia |
| Hyperglycemia | R73.09, E11.65 | Elevated blood glucose / hyperglycemia with diabetes |

##### **6.5 Musculoskeletal Adverse Events**

| **Adverse Event** | **ICD-10 Code(s)** | **Description** |
| --- | --- | --- |
| Myalgia | M79.1 | Non-specific muscle pain |
| Myositis | M60 | Inflammatory myopathy |
| Rhabdomyolysis | M62.82 | Severe skeletal muscle breakdown |

##### **6.6 Dermatologic Adverse Events**

| **Adverse Event** | **ICD-10 Code(s)** | **Description** |
| --- | --- | --- |
| Urticaria / hives | L50 | Allergic or idiopathic urticaria |
| Rash (non-specific) | R21 | Non-specific skin eruption |
| Pruritus | L29, L29.9 | Generalized or localized itching |
| Photosensitivity | L56, L56.8 | Photoallergic or phototoxic reaction |
| Alopecia | L65, L65.9 | Non-scarring hair loss |

##### **6.7 Respiratory Adverse Events**

| **Adverse Event** | **ICD-10 Code(s)** | **Description** |
| --- | --- | --- |
| Dyspnea | R06.0, R06.00, R06.09 | Shortness of breath |
| Cough | R05 | Non-specific cough |

##### **6.8 Ophthalmic, Genitourinary, Hepatic, and Hypersensitivity Adverse Events**

| **Adverse Event** | **ICD-10 Code(s)** | **Description** |
| --- | --- | --- |
| Blurred vision | H53.8, H53.10 | Visual disturbance / blurring |
| Urinary retention | R33, R33.9 | Incomplete bladder emptying |
| Drug-induced liver injury (DILI) | K71, K71.6 | Toxic hepatopathy / drug-induced hepatitis |
| Hypersensitivity reactions (broad) | J39.3, J67, M31.0, T78.1, T78.40, T80.6 | Broad allergic and immunologic hypersensitivity |
| Angioedema | T78.3 | Allergic swelling of subcutaneous tissue |
| Anaphylaxis | T78.0, T78.2, T80.5, T88.6 | Severe systemic allergic reaction |

#### 7. Summary of Data Types and Measurement Approach

All variables were measured across every applicable time window (1 baseline + 3 outcome windows). The following data types were captured:

| **Domain** | **Variable Type** | **Measurement Approach** |
| --- | --- | --- |
| Elixhauser comorbidities (28 conditions) | Binary (present/absent) | Any ICD-10/ICD-9 diagnosis code within the time window |
| Healthcare utilization (14 categories) | Binary + continuous (count or LOS) | CPT codes and/or visit type flags; count of encounters or duration in days |
| Laboratory values (30 measures) | Continuous (last observed value) | LOINC-coded lab results filtered to plausible range; last observation per window |
| Laboratory/vital thresholds (57 cutpoints) | Binary (above/below threshold) | Plausibility-filtered LOINC values dichotomized at clinically meaningful cutpoints |
| Change from baseline (Δ, 30 measures) | Continuous (delta) | Outcome window last value minus baseline last value; range-bounded |
| Adverse events (43 categories) | Binary (present/absent) | ICD-10 diagnosis codes within the time window |

### Part B — Statistical Methods

#### 8. Adjusted Estimator: Estimation Details

The adjusted estimator is the scalable collaborative TMLE of Ju et al. ^1^ (Figure S2). Both nuisance functions, the outcome regression (Q) and the treatment/propensity model (g), are estimated with cross-validated gradient-boosted trees (XGBoost, 50 boosting rounds) under 5-fold cross-fitting. Candidate confounders comprise the full baseline covariate set (demographics, the 28 Elixhauser comorbidities, baseline laboratory and vital-sign measures, and healthcare-utilization indicators measured in the pre-index window); the 185 baseline features used for effect-measure modification are a subset used only for subgroup stratification, not a separate adjustment set. Before the collaborative search, a double-selection screen restricts candidates to covariates predictive of both treatment and outcome, excluding pure instruments. Covariates are then ranked once by their approximate contribution to the bias term (XGBoost gain on arm-demeaned outcome residuals), and nested propensity models (the ordered prefixes of this ranking, up to 25 candidates) are evaluated by cross-validated negative log-likelihood; the prefix minimizing held-out loss is selected, with patience-based early stopping after three consecutive non-improving prefixes. Estimated propensity scores are truncated at 0.025 (bounded clever covariate). Standard errors are obtained from the estimated efficient influence function, and Wald-type 95% confidence intervals are constructed. For multi-arm comparisons the treatment model is multinomial, and treatment-specific means are estimated for each arm with pairwise contrasts against a common reference arm.

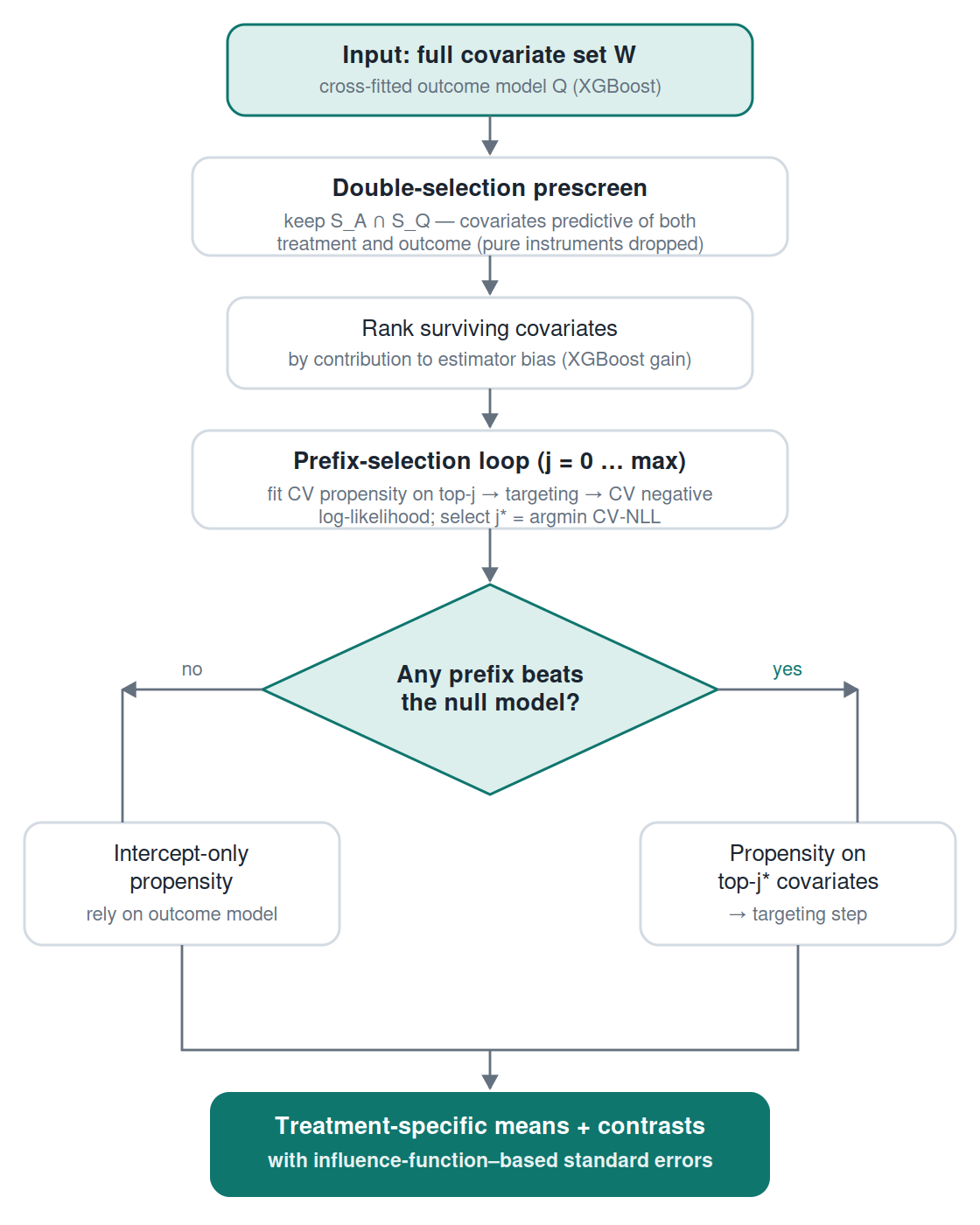

Figure S2. Collaborative TMLE covariate-selection procedure. Flowchart of the adjusted estimator: a double-selection prescreen retains covariates predictive of both treatment and outcome; survivors are ranked by their approximate contribution to the estimator's bias term; nested propensity models over ordered prefixes of the ranking are evaluated by cross-validated negative log-likelihood, and the loss-minimizing prefix is selected (with patience-based early stopping). If no prefix improves on the null, an intercept-only propensity model is used, relying on the outcome regression. The targeting step yields treatment-specific means and pairwise contrasts with influence-function–based standard errors.

Missing data. Analyses of continuous outcomes are complete-case within each follow-up window: patients without a measured value in a window are excluded from that window's analysis, which is valid under missingness at random given the covariates in the outcome model ^2^. For binary and discrete outcomes, when no qualifying event is observed in the source data the outcome is coded 0 rather than treated as missing. Because as-treated follow-up is censored at discontinuation, worse-tolerated arms accrue less observation time; adverse-event risk differences under this coding should be read as on-treatment contrasts rather than incidence contrasts.

Downsampling. To bound computational cost in very large cohorts, each treatment arm was randomly downsampled without replacement to a common per-arm maximum of 5,000 patients before estimation, and both the adjusted and unadjusted analyses used this common downsampled population. Because the collaborative covariate selection depends on the sampled data, the selected adjustment set can vary with the random seed.

Positivity and E-values. Estimated propensity scores were truncated at 0.025 (bounded clever covariate), and per-arm positivity and effective-sample-size diagnostics were retained. For each effect estimate we computed an E-value — the minimum association, on the risk-ratio scale, that an unmeasured confounder would need with both treatment and outcome to explain away the observed effect. Here the E-value is derived from the standardized effect size (a standardized mean difference mapped to an approximate risk ratio) rather than a directly modeled risk ratio; because this mapping stacks approximations across binary and continuous outcomes, the resulting E-values are approximate robustness indices rather than strictly comparable quantities.

Residual-confounding (causal-gap) sensitivity. When both the adjusted and unadjusted estimators completed for an outcome, we additionally ran a residual-confounding sensitivity analysis following Luedtke, Díaz, and van der Laan ^3^ that quantifies how large a departure from conditional exchangeability — a bounded causal gap between the adjusted estimate and the true effect — would be required to overturn a finding.

#### 9. Unadjusted Sensitivity Analysis

As a sensitivity analysis complementing the primary confounding-adjusted collaborative TMLE (Ju C-TMLE) estimates, we additionally computed unadjusted treatment comparisons for every outcome. Unadjusted estimates make no modeling assumptions about confounding structure: they reflect differences between treatment groups as they exist in the data, combining any treatment effect with differences in patient characteristics between arms. They serve as a transparent, reproducible benchmark, and substantial divergence between the adjusted and unadjusted estimates identifies the scenarios in which measured confounding is most consequential.

##### **9.1 Treatment-Specific Means**

For each treatment arm we computed the unadjusted mean outcome. For continuous and discrete outcomes, the standard error was the sample standard deviation divided by the square root of the arm sample size. For binary outcomes, the treatment-specific mean is the event proportion p and its standard error is the binomial standard error, the square root of p(1 − p)/n.

##### **9.2 Effect Estimates**

Pairwise effect estimates (each non-reference arm versus the reference arm) were obtained by regressing the outcome on the treatment indicator(s) by ordinary least squares, using the same approach for continuous, discrete, and binary outcomes; this yields a mean difference for continuous and discrete outcomes and a risk difference for binary outcomes (equivalently, a linear probability model). Standard errors were heteroskedasticity-consistent (HC3), and Wald-type 95% confidence intervals were constructed for each estimate.

##### **9.3 Effect-Measure Modification**

To assess effect-measure modification, the same unadjusted regression was applied within each stratum defined by a baseline modifier, producing stratum-specific mean or risk differences.

##### **9.4 Missing Data and Multiplicity**

All continuous-outcome analyses were complete-case: patients missing an outcome value in a given window were excluded from that window's analysis. For binary and discrete outcomes, missing outcomes were set to 0. Consistent with the primary analysis, Benjamini–Hochberg false-discovery-rate–adjusted p-values are reported alongside unadjusted p-values; no family-wise correction is imposed, and results are treated as exploratory and interpreted in aggregate and in clinical context.

#### 10. Effect-Measure Modification Test

To screen for effect-measure modification we test, for each candidate modifier V, the null hypothesis that the between-arm treatment contrast does not vary with V. Under this null the within-arm effect of V (the difference in an arm's treatment-specific mean between strata, EMM_k = TSM_k(V=1) − TSM_k(V=0)) is identical across arms (V shifts all arms additively, leaving between-arm contrasts unchanged). The statistic is a precision-weighted homogeneity chi-square on the arm-specific V-effects, χ² = Σ_k w_k (EMM_k − EMM_pool)², with weights w_k = 1/SE(EMM_k)² and EMM_pool = Σ_k w_k EMM_k / Σ_k w_k, on (number of arms − 1) degrees of freedom. The arm-specific V-effects are treated as independent: to first order under cross-fitting, the influence function for arm k is supported only on patients assigned to arm k (through the treatment indicator in the clever covariate), so cross-fold influence contributions for distinct arms are orthogonal and the off-diagonal covariances vanish. This first-order argument neglects higher-order coupling through the shared nuisance estimators; formal calibration of the test's type-I error and coverage is part of the companion validation. Continuous modifiers are dichotomized at the sample median for this screen, and the test is used to rank and flag candidate modifiers rather than as a confirmatory hypothesis test.

### Part C — Automation & Scope

#### 11. Automated Narrative Summaries — LLM Configuration

*Narrative summaries were produced by a hybrid deterministic/LLM pipeline. All quantitative content—effect estimates, confidence intervals, sample sizes, and significance flags—is rendered deterministically from the estimate tables; large language models are used only for language tasks (expanding encoded outcome and subgroup names into plain English and drafting the summary paragraph) and never to compute, select, or modify any estimate.*

*Summaries were generated with Gemini 3.1 Pro (gemini-3.1-pro-preview) at temperature 0 (deterministic decoding), a 4,096-token output limit, and up to three retries; Claude (claude-opus-4-6, claude-sonnet-4-6) and Gemini 2.5 Pro are supported as alternatives. The pipeline runs in three phases: (A) an LLM phase that classifies each outcome name (plain-English label, clinical domain, binary/continuous type, and change-from-baseline flag) and expands encoded strata-variable names into prose noun phrases, with results cached across runs; (B) a deterministic phase that assembles the quantitative Results section, organized by clinical domain and significance and rendered as templated sentences with effect sizes, confidence intervals, and sample sizes; and (C) a final LLM phase that writes only the summary prose from the deterministic Phase B text.*

#### 12. Full List of Clinical Scenarios

*The 135 treatment-comparison scenarios analyzed in this study are listed below, corresponding to the domain-level summary in Table 1 of the main text.*

*1. Atrial Fibrillation on Rate Control AddOn Antiarrhythmics vs Catheter Ablation*

*2. Acute Myocardial Infarction Management PCI vs Fibrinolytics*

*3. ADHD Initial Therapy Amphetamine Salt Double Bead vs Triple Bead Mydayis vs Easy Swallow Adzenys vs Methylphenidate ER*

*4. Atopic Dermatitis Initial Therapy Triamcinolone Topical vs Crisaborole Topical*

*5. Dry Eye Syndrome Bilateral Initial Therapy Cyclosporine Ophthalmic vs Lifitegrast Ophthalmic*

*6. Endometriosis Initial Therapy Combined Oral Contraceptives vs GnRH Agonists vs GnRH Antagonists*

*7. Fibromyalgia Initial Therapy Pregabalin vs Duloxetine vs Milnacipran*

*8. Focal Seizures Initial Therapy Levetiracetam vs Lamotrigine DISC*

*9. Gout Prophylaxis Initial Therapy Allopurinol vs Febuxostat*

*10. Herpes Zoster Initial Therapy Acyclovir vs Valacyclovir*

*11. HFrEF on ACE Inhibitor or ARB AddOn SGLT2I vs MRA vs Entresto*

*12. HIV Initial Therapy Biktarvy vs Triumeq*

*13. Insomnia Initial Therapy Trazodone vs Zolpidem vs Suvorexant*

*14. Obesity BMI 35 or Greater Initial Therapy Semaglutide High Dose vs Tirzepatide H*

*15. Parkinsons Disease Initial Therapy Carbidopa Levodopa Combination vs Pramipexol DISC*

*16. PCOS Initial Therapy Combined Oral Contraceptives vs Metformin*

*17. T2DM On Metformin Poor Control AddOn Sulfonylurea vs DPP4I vs Insulin vs GLP1RA vs Tirzepatide vs SGLT2I*

*18. Ventricular Tachycardia on Beta Blockers AddOn Amiodarone vs VT Catheter Ablation vs Sotalol*

*19. Cerebrovascular Disease With Atherosclerosis Antiplatelet Therapy Clopidogrel vs Ticagrelor*

*20. Overactive Bladder Initial Therapy Oxybutynin Extended Release vs Mirabegron*

*21. Chronic Myeloid Leukemia Initial Therapy Imatinib vs Dasatinib*

*22. Community Acquired Pneumonia Outpatient Low Risk Amoxicillin vs Azithromycin vs*

*23. Alopecia Areata JAK Inhibitor Comparison Baricitinib vs Ritlecitinib*

*24. AFib Rate Control vs Rhythm Control Pharmacological Strategy*

*25. Early Catheter Ablation vs Antiarrhythmic Drug Therapy as First Line Rhythm Cont*

*26. Warfarin vs NOAC Stroke DVT PE Outcomes in Atrial Fibrillation*

*27. ASCVD High-Intensity Statin vs Ezetimibe First-Line Therapy*

*28. Type 2 Diabetes Metformin-Treated Add-On SGLT2i vs DPP-4i*

*29. Carotid Endarterectomy vs Carotid Artery Stenting in Symptomatic Carotid Stenosi*

*30. Inpatient Rehabilitation Facility vs Home Based Rehabilitation Post Stroke*

*31. In Center Hemodialysis vs Home Dialysis in CKD Dialysis Dependent Patients*

*32. AV Fistula vs AV Graft for Hemodialysis Access in CKD Dialysis Patients*

*33. Effectiveness of Osteoperosis treatment with Romosozumab vs Teriparatide*

*34. Alpha2 Delta Ligands vs Ropinirole in Restless Legs Syndrome*

*35. Effectivness of pulmonary arterial hypertension treatment with Sildenafil vs Selexipag or Riociguat*

*36. Antiplatelet P2Y12 Inhibitors vs Apixaban vs Rivaroxaban vs Warfarin in Cerebrov*

*37. CGRP Monoclonal Antibody vs Botox for Chronic Migraine Prevention*

*38. CKD Anemia Epoetin Alfa vs Darbepoetin Alfa ESA Head to Head*

*39. Donepezil Plus Memantine vs Donepezil Alone vs Memantine Alone vs Rivastigmine i*

*40. Spinal Cord Stimulation vs Medical Management for Painful Diabetic Peripheral Ne*

*41. Preemptive Kidney Transplant vs Dialysis First in CKD*

*42. Peritoneal Dialysis vs Hemodialysis in CKD Dialysis Dependent Patients*

*43. Oral CGRP Antagonists vs Subcutaneous CGRP Antagonists vs Botulinum Toxin for Mi*

*44. Modafinil vs Solriamfetol for Narcolepsy Wake Promoting Agent Comparison*

*45. Modafinil vs Armodafinil for Narcolepsy Type 2 Wake Promoting Efficacy and Safet*

*46. Gabapentinoids vs Amitriptyline vs Duloxetine vs Opioids for Peripheral Neuropat*

*47. Drug Resistant Epilepsy Surgery vs Continued AED Therapy*

*48. Carotid Revascularization vs Medical Management in Asymptomatic Carotid Stenosis*

*49. Rheumatoid Arthritis Initial Biologic Therapy TNF Inhibitor vs JAK Inhibitor*

*50. Stimulant vs Non-Stimulant ADHD Pharmacotherapy Comparative Effectiveness in Adu*

*51. Rheumatoid Arthritis First Line Therapy TNF Inhibitors vs Non TNF Biologics*

*52. Psoriatic Arthritis First Line Therapy TNF Inhibitors vs Non TNF Biologics*

*53. Ankylosing spondylitis First Line Therapy TNF Inhibitors vs Non TNF Biologics*

*54. Crohns Disease First Line Therapy TNF Inhibitors vs Non TNF Biologics*

*55. Ulcerative Colitis First Line Therapy TNF Inhibitors vs Non TNF Biologics*

*56. Plaque Psoriasis First Line Therapy TNF Inhibitors vs Non TNF Biologics*

*57. Juvenile ideopathic arthritis First Line Therapy TNF Inhibitors vs Non TNF Biolo*

*58. COPD Maintenance Therapy LAMA vs LABA Plus ICS vs LAMA Plus LABA*

*59. COPD Pulmonary Rehabilitation vs Usual Care*

*60. Moderate to Severe Asthma Biologic Therapy Omalizumab vs Mepolizumab vs Dupilumab*

*61. Asthma SMART ICS Formoterol Maintenance and Reliever vs ICS plus SABA PRN*

*62. Severe Asthma Biologic Omalizumab vs Mepolizumab vs Dupilumab*

*63. Acne Vulgaris Topical Retinoid Tretinoin vs Adapalene*

*64. Benign Prostatic Hyperplasia Second Line Therapy after Alpha-1 Blockers Finasteride vs Dutasteride vs Tadalafil*

*65. Iron Deficiency Anemia Oral Iron vs Intravenous Iron*

*66. Localized Prostate Cancer Radical Prostatectomy vs Radiation Therapy*

*67. Cancer Associated Thrombosis LMWH vs DOAC*

*68. Metastatic Prostate Cancer ADT plus ARPI vs ADT Alone*

*69. Early Breast Cancer Breast Conserving Surgery vs Mastectomy*

*70. Eosinophilic Esophagitis Swallowed Topical Corticosteroid vs Proton Pump Inhibitor*

*71. Hemorrhoids Rubber Band Ligation vs Hemorrhoidectomy*

*72. SSRI vs SNRI vs Buspirone Outcomes in Anxiety Generalized Anxiety Disorder*

*73. Buprenorphine Naloxone vs Methadone vs Naltrexone for Opioid Use Disorder*

*74. BPH Finasteride vs Dutasteride 5-ARI add-on to Alpha-Blocker*

*75. Cabergoline vs Bromocriptine in Hyperprolactinemia Prolactinoma*

*76. Corticosteroid vs Methotrexate vs Anti-TNF in Pulmonary Sarcoidosis*

*77. Osteoarthritis Knee Intraarticular Corticosteroid vs Hyaluronic Acid Injection*

*78. Oral vs IV Antibiotics for Uncomplicated Cellulitis SSTI*

*79. TMS vs Medication Switch Outcomes in Treatment-Resistant Major Depressive Disorder*

*80. Chronic Urticaria Initial Therapy Dupilumab vs Omalizumab*

*81. Chronic sinustitis dupilumab vs Omalizumab*

*82. Fidaxomicin vs Oral Vancomycin for Initial Clostridioides difficile Infection CDI*

*83. Chronic Hepatitis B Tenofovir vs Entecavir*

*84. Hepatic Encephalopathy Lactulose vs Rifaximin*

*85. Rosacea Oral Doxycycline vs Topical Metronidazole vs Topical Ivermectin*

*86. Alpha-Blocker Initial Therapy for BPH Tamsulosin vs Doxazosin vs Terazosin vs Alfuzosin*

*87. CBT vs Pharmacotherapy Outcomes in Anxiety and Generalized Anxiety Disorder*

*88. SSRI vs SNRI Initial Therapy in Major Depressive Disorder*

*89. Chronic Hepatitis B Tenofovir Disoproxil vs Tenofovir Alafenamide*

*90. Ulcerative Colitis Biologic Therapy TNF Inhibitor vs Vedolizumab vs Ustekinumab*

*91. Open Angle Glaucoma First Line Selective Laser Trabeculoplasty vs Topical IOP Lowering Eye Drops*

*92. Onychomycosis Oral Terbinafine vs Topical Efinaconazole*

*93. Nintedanib vs Pirfenidone in Idiopathic Pulmonary Fibrosis*

*94. Ankylosing Spondylitis Biologic DMARD Comparison TNF Inhibitors vs IL17 Inhibitors*

*95. IV Iron vs Dopamine Agonist for Restless Legs Syndrome Iron Repletion*

*96. Medications for Opioid Use Disorder Alone vs MOUD Plus Psychosocial Intervention*

*97. Erectile Dysfunction PDE5 Inhibitor Sildenafil vs Tadalafil*

*98. Endometriosis Laparoscopic Excision vs Medical Management Dienogest*

*99. Obstructive Sleep Apnea CPAP vs Oral Appliance Mandibular Advancement Therapy*

*100. Venous Thromboembolism Acute Treatment DOAC vs Warfarin*

*101. Menopausal Vasomotor Symptoms Hormone Therapy vs Fezolinetant vs SSRI*

*102. Uterine Fibroids Uterine Artery Embolization vs Myomectomy vs Hysterectomy*

*103. Post Traumatic Stress Disorder SSRI SNRI Pharmacotherapy vs Trauma Focused Psychotherapy*

*104. Panic Disorder SSRI vs Cognitive Behavioral Therapy*

*105. Social Anxiety Disorder SSRI vs Cognitive Behavioral Therapy*

*106. Treatment Resistant Depression Esketamine vs Aripiprazole vs Lithium Augmentation*

*107. Obstructive Sleep Apnea Tirzepatide vs CPAP Device Therapy*

*108. Tobacco Use Disorder Smoking Cessation Varenicline vs Bupropion vs Nicotine Replacement Therapy*

*109. Anorexia Nervosa Family-Based Therapy vs Individual Psychotherapy in Adolescents*

*110. Sjogren Syndrome Oral Secretagogue Pilocarpine vs Cevimeline*

*111. Propranolol vs Primidone Outcomes in Essential Tremor*

*112. Insulin vs Metformin in Gestational Diabetes*

*113. Apixaban vs Rivaroxaban vs Dabigatran in Atrial Fibrillation*

*114. CBT-I vs Pharmacotherapy Outcomes in Insomnia*

*115. Oseltamivir vs Baloxavir Marboxil for Acute Influenza*

*116. Chronic Spontaneous Urticaria Omalizumab vs High-Dose Antihistamine*

*117. DPP-4 Inhibitors vs Sulfonylureas as Second-Line Therapy in Type 2 Diabetes*

*118. Osteoporosis Oral Bisphosphonate vs IV Zoledronic Acid*

*119. Menorrhagia Tranexamic Acid vs Norethindrone vs Combined Oral Contraceptive for Heavy Menstrual Bleeding*

*120. Nirmatrelvir-Ritonavir vs Molnupiravir Outpatient High-Risk COVID-19 Outcomes*

*121. Finerenone vs SGLT2 Inhibitor for Disease Modification in Diabetic CKD*

*122. Ketoconazole vs Topical Corticosteroid Outcomes in Seborrheic Dermatitis*

*123. Multiple Myeloma Transplant-Eligible Dara-VRd Quadruplet vs VRd Triplet*

*124. Multiple Myeloma Transplant-Ineligible Dara-Rd vs VRd*

*125. Endovascular Repair EVAR vs Open Surgical Repair for Abdominal Aortic Aneurysm*

*126. Sofosbuvir-Velpatasvir vs Glecaprevir-Pibrentasvir Outcomes in Hepatitis C*

*127. Patiromer vs Sodium Zirconium Cyclosilicate Outcomes in Chronic Hyperkalemia*

*128. Thiazide vs ACE Inhibitor vs Calcium Channel Blocker Initial Therapy Outcomes in Hypertension ALLHAT*

*129. Resistant Hypertension Spironolactone Add-On vs Other 4th-Line Antihypertensive Agent*

*130. Anti-VEGF vs Intravitreal Dexamethasone Implant Outcomes in Retinal Vein Occlusion*

*131. Bisphosphonate vs Denosumab in Osteoporosis*

*132. COPD Maintenance LAMA vs LABA vs LAMA-LABA Dual Therapy*

*133. Roflumilast vs Azithromycin for COPD Frequent Exacerbators*

*134. Levodopa vs Pramipexole vs Rasagiline in Parkinson Disease Initial Therapy*

*135. Sevelamer vs Lanthanum vs Ferric Citrate in Hyperphosphatemia ESRD*

*Abbreviations: ACC, American College of Cardiology; ACP, advance care planning; ACR, albumin-to-creatinine ratio; ADA, American Diabetes Association; AHA, American Heart Association; ALT, alanine aminotransferase; AST, aspartate aminotransferase; BMP, basic metabolic panel; BMI, body mass index; BUN, blood urea nitrogen; CBC, complete blood count; CHF, congestive heart failure; CKD, chronic kidney disease; CMP, comprehensive metabolic panel; CMS, Centers for Medicare & Medicaid Services; CPT, Current Procedural Terminology; CRP, C-reactive protein; CT, computed tomography; CV, cardiovascular; DBP, diastolic blood pressure; DEXA, dual-energy X-ray absorptiometry; DILI, drug-induced liver injury; ED, emergency department; eGFR, estimated glomerular filtration rate; EHR, electronic health record; E/M, evaluation and management; GGT, gamma-glutamyl transferase; GI, gastrointestinal; HbA1c, hemoglobin A1c; HDL, high-density lipoprotein; hs-CRP, high-sensitivity C-reactive protein; HTN, hypertension; IBS, irritable bowel syndrome; ICD-9, International Classification of Diseases, Ninth Revision; ICD-10, International Classification of Diseases, Tenth Revision; ICU, intensive care unit; INR, International Normalized Ratio; LDL, low-density lipoprotein; LOINC, Logical Observation Identifiers Names and Codes; LOS, length of stay; MDD, major depressive disorder; MRI, magnetic resonance imaging; NOS, not otherwise specified; NT-proBNP, N-terminal pro-B-type natriuretic peptide; OT, occupational therapy; PT, physical therapy; PT-INR, prothrombin time–International Normalized Ratio; RWD, real-world data; SBP, systolic blood pressure; SGOT, serum glutamic-oxaloacetic transaminase; SGPT, serum glutamic-pyruvic transaminase; SNF, skilled nursing facility; TCM, transitional care management; TG, triglycerides; TSH, thyroid-stimulating hormone; ULN, upper limit of normal; WBC, white blood cells.*

#### References

*1.	Ju C, Gruber S, Lendle SD, others. Scalable collaborative targeted learning for high-dimensional data. Statistical Methods in Medical Research 2019;28(2):532–54.*

*2.	Little RJA, Rubin DB. Statistical Analysis with Missing Data. 3rd ed. Hoboken, NJ: Wiley; 2019.*

*3.	Luedtke AR, Díaz I, van der Laan MJ. The statistics of sensitivity analyses [Internet]. U.C. Berkeley Division of Biostatistics; 2015. Available from: https://biostats.bepress.com/ucbbiostat/paper341/*
